# Leveraging genetic data to elucidate the relationship between Covid-19 and ischemic stroke

**DOI:** 10.1101/2021.02.25.21252441

**Authors:** Verena Zuber, Alan Cameron, Evangelos P. Myserlis, Leonardo Bottolo, Israel Fernandez-Cadenas, Stephen Burgess, Christopher D. Anderson, Jesse Dawson, Dipender Gill

## Abstract

**Background:** The relationship between coronavirus disease 2019 (Covid-19) and ischemic stroke is poorly defined. We aimed to leverage genetic data to investigate reported associations.

**Methods:** Genetic association estimates for liability to Covid-19 and cardiovascular traits were obtained from large-scale consortia. Analyses primarily focused on critical Covid-19, defined as hospitalization with Covid-19 requiring respiratory support or resulting in death. Cross-trait linkage disequilibrium score regression was used to estimate genetic correlations of critical Covid-19 with ischemic stroke, other related cardiovascular outcomes, and risk factors common to both Covid-19 and cardiovascular disease (body mass index, smoking and chronic inflammation, estimated using C-reactive protein). Mendelian randomization analysis was performed to investigate whether liability to critical Covid-19 was associated with increased risk of any of the cardiovascular outcomes for which genetic correlation was identified.

**Results:** There was evidence of genetic correlation between critical Covid-19 and ischemic stroke (r_g_=0.29, FDR *p*-value=4.65×10^−3^), body mass index (r_g_=0.21, FDR-*p*-value*=*6.26×10^−6^) and C-reactive protein (r_g_=0.20, FDR-*p*-value=1.35×10^−4^), but none of the other considered traits. In Mendelian randomization analysis, liability to critical Covid-19 was associated with increased risk of ischemic stroke (odds ratio [OR] per logOR increase in genetically predicted critical Covid-19 liability 1.03, 95% confidence interval 1.00-1.06, *p*-value=0.03). Similar estimates were obtained when considering ischemic stroke subtypes. Consistent estimates were also obtained when performing statistical sensitivity analyses more robust to the inclusion of pleiotropic variants, including multivariable Mendelian randomization analyses adjusting for potential genetic confounding through body mass index, smoking and chronic inflammation. There was no evidence to suggest that genetic liability to ischemic stroke increased the risk of critical Covid-19.

**Conclusions:** These data support that liability to critical Covid-19 is associated with an increased risk of ischemic stroke. The host response predisposing to severe Covid-19 is likely to increase the risk of ischemic stroke, independent of other potentially mitigating risk factors.

## Introduction

Severe acute respiratory syndrome coronavirus 2 (SARS-CoV-2) infection is the cause of the coronavirus disease 2019 (Covid-19) pandemic that has resulted in a health crisis of unprecedented magnitude.^1, 2^ While much of the disease burden relates to respiratory failure and sepsis, some studies suggest an increased risk of ischemic stroke.^3–6^ This has been estimated to be seven times greater than in influenza infection,^3^ with up to 5% of people with severe Covid-19 suffering stroke.^5^ Strokes that occur in individuals with Covid-19 are more severe, have poorer outcomes and higher mortality rates than in those without Covid-19, despite similar acute managment.^6, 7^ Indeed, almost two-fifths of people with Covid-19 who develop stroke consequently die.^8^ However, some studies do not support an increased risk of stroke in individuals with Covid-19.^9, 10^ Obtaining unbiased estimates for the risk of stroke in people with Covid-19 is challenging due to difficulty diagnosing mild Covid-19 and an overall reduction in the rate of admission to hospital with stroke, and minor stroke in particular, during the pandemic.^9, 11^ Furthermore, observational studies investigating the association between Covid-19 and stroke are vulnerable to potential confounding and reverse causation.^3–6^ For example, there are common risk factors for severe Covid-19 and stroke, such as obesity and smoking^12^. Similarly, patients with acute stroke have a dampened immune response and may be more susceptible to severe Covid-19.^13^

Leverage of genetic data can help overcome some of these issues. Cross-trait linkage disequilibrium score regression (LDSC) can be used to estimate the genetic correlation between traits. Mendelian randomization (MR) can be employed to investigate whether genetic variants predicting an exposure (such as Covid-19) also associate with risk of an outcome (such as ischemic stroke).^14^ There are numerous plausible mechanisms by which Covid-19 may be increasing ischemic stroke risk. Covid-19 can trigger a cytokine storm with upregulation of pro-inflammatory signaling and endothelial dysfunction that predisposes to a hypercoagulable state and can lead to thromboembolic events.^15^ Indeed, Covid-19 also appears to promote the development of other cardiovascular disorders including myocardial injury, myocardial ischemia, arrhythmias, heart failure and venous thromboembolism.^15^ Furthermore, pre-existing cardiovascular disease (CVD) is associated with high mortality in people with Covid-19, which has raised the possibility of a bidirectional interaction between Covid-19 and the cardiovascular system.^15^ MR analyses can also allow the exploration of such bidirectional relationships.

Elucidating the relationship between Covid-19 and risk of ischemic stroke could prove important for optimizing prevention and treatment strategies. With this in mind, we performed cross-trait LDSC to investigate whether there is a genetic correlation between Covid-19 and ischemic stroke, and followed this up with MR analyses to investigate whether any such statistically significant correlation might be explained by liability to Covid-19 being associated with increased risk of ischemic stroke.

## Methods

All genetic association data used in this work are publicly accessible. Appropriate patient consent and ethical approval had been obtained in the original studies from which they were obtained (**Supplementary Table 1**). Statistical code related to the analyses performed in the current study is freely available from Github (https://github.com/verena-zuber/covid19_and_stroke).

### Study overview

First, we performed cross-trait LDSC to estimate genetic correlations for Covid-19 with ischemic stroke, other related CVDs, and risk factors common to both Covid-19 and CVD. Second, for CVD outcomes that showed evidence of genetic correlation with Covid-19, MR analysis was performed to investigate whether liability to Covid-19 was also associated with these outcomes. Finally, bidirectional MR was carried out to investigate potential reverse associations, i.e. whether genetic liability to the CVD outcome was also associated with increased risk of Covid-19.

### Exposure definitions and genetic association estimates for Covid-19

Genetic association estimates for Covid-19 were obtained from release 5 of the Covid-19 host genetics consortium.^16^ In our analysis we focused on the most severe definition of Covid-19 available, where a critical case is defined as an individual who was hospitalized with laboratory confirmed SARS-CoV-2 infection and required respiratory support or died. Genetic associations were derived from 5,101 cases and 1,383,241 controls from the general population. Hospital admission and requiring respiratory support or death is a proxy for disease severity and is preferred here over other case definitions which are solely based on a positive Covid-19 test result. Previous studies have shown that bias may impact analyses identifying cases based on likelihood of testing for SARS-CoV-2 infection, because participants being tested for SARS-CoV-2 infection are selected for a wide range of genetic, behavioral and demographic traits.^9^

Results based on other Covid-19 definitions from the Covid-19 host genetics consortium were performed as further sensitivity analysis. Firstly, we compared individuals with laboratory confirmed SARS-CoV-2 infection who had been hospitalized (cases) versus individuals with laboratory confirmed SARS-CoV-2 infection who did not require hospitalization (4,829 cases and 11,816 controls). Secondly, we compared individuals with laboratory confirmed SARS-CoV-2 infection who had been hospitalized (cases) versus the general population (9,986 cases and 1,877,672 controls). The final definition was based on individuals with reported Covid-19 (laboratory confirmed, physician-reported or self-reported; cases) versus controls from the general population (38,984 cases and 1,644,784 controls).

### Outcomes

#### Ischemic stroke

The primary outcome was any ischemic stroke (34,217 cases). In secondary, hypothesis-generating analyses, stroke subtypes were further explored as large artery stroke (LAS, 4,373 cases), cardioembolic stroke (CES, 7,193 cases) and small vessel stroke (SVS, 5,386 cases).^17^ The common control pool included 406,111 individuals. Genetic association data were derived from the MEGASTROKE consortium.^18^

#### Related cardiovascular disease outcomes

We considered other CVD outcomes related to ischemic stroke in their pathophysiology. These were coronary artery disease (including myocardial infarction, acute coronary syndrome, chronic stable angina or >50% coronary artery stenosis), heart failure and atrial fibrillation. Genetic associations with risk for coronary artery disease were measured on 60,801 cases and 123,504 controls and taken from the Coronary ARtery DIsease Genome wide Replication and Meta-analysis (CARDIOGRAM) plus The Coronary Artery Disease (C4D) consortium (CARDIoGRAMplusC4D),^19^ for heart failure were measured on 47,309 cases and 930,014 controls and taken from the HEart failuRe Molecular Epidemiology for therapeutic targetS (HERMES) consortium^20^ and for atrial fibrillation were measured on 65,446 cases and 522,744 controls and taken from a transethnic meta-analysis.^21^

### Risk factors related to both Covid-19 and cardiovascular disease

To investigate whether any genetic correlation between critical Covid-19 and the CVD outcomes was related to confounding factors, we further considered common risk factors to both, including obesity, smoking and chronic inflammation.^22–25^ Genetic association estimates to proxy these traits were taken from a genome-wide association study (GWAS) on body mass index (BMI) measured on 694,649 subjects,^26^ lifetime smoking index measured on 462,690 subjects,^27^ and C-reactive protein (CRP) measured on 361,194 individuals in UK Biobank ^28^.

### Statistical analyses

#### Cross-trait linkage disequilibrium score regression

We performed LDSC to estimate the genetic correlation (r_g_) of critical Covid-19 with the primary outcome ischemic stroke, and secondary outcomes coronary artery disease, heart failure and atrial fibrillation, using GWAS summary statistics data.^29,30^ We also estimated correlation with possible genetic confounders, including BMI, lifetime smoking index and CRP. We restricted our analyses to HapMap 3 single-nucleotide polymorphisms (SNPs), which are known to be well-imputed across most studies and utilized the pre-computed European LD-scores estimated using the 1000G reference panel, provided by the LDSC creators. For each set of summary statistics, the SNP-specific sample size information was used. If not available, we assumed that all SNPs had the same sample size for that trait, defined as the total sample size for continuous phenotypes or as the sum of cases and controls for case/control phenotypes. By default, LDSC also removed variants that were duplicate, strand-ambiguous, not SNPs (e.g. indels), with *p*-values not between 0 and 1, with alleles that did not match with the 1000G reference panel, and with low effective sample size or not included in all studies of a GWAS meta-analysis (if such information was available) for traits with no effective sample size information. After estimation of the genetic correlation across all phenotypes, we corrected for multiple hypothesis testing using the Benjamini and Hochberg false discovery rate (FDR).^31^ FDR-corrected *p*-values <0.05 were considered statistically significant.

#### Mendelian randomization analyses

##### Genetic variants used as instrumental variables

Genetic variants were selected based on associations with critical Covid-19. In our main analysis, we selected uncorrelated genetic variants (clumped at correlation threshold *r*^*2*^*<0*.*01*) at *p*-value<5×10^−6^. In sensitivity analyses, we applied a more stringent threshold and considered only genome-wide significant genetic variants (*p*-value<5×10^−8^).

##### Main analysis

For CVD outcomes that showed evidence of genetic correlation with critical Covid-19 in LDSC, MR analysis was performed to estimate the association of genetically predicted liability to critical Covid-19 with that outcome using the random effects two-sample inverse-variance weighted (IVW) method.^32^ The IVW estimate can be biased by pleiotropy when a genetic variant associates the outcome (e.g., ischemic stroke) via a pathway other than through the exposure (i.e., liability to critical Covid-19). Pleiotropy can cause heterogeneity in the MR estimates obtained by different variants employed as instruments, which was assessed using the *Q-*statistic and the respective heterogeneity *p-*value.^33^ A Mendelian randomization estimate with *p-*value<0.05 for the main IVW analysis was deemed to represent supportive evidence, given that MR was only performed to follow up positive LDSC findings.

##### Sensitivity Analyses

We performed sensitivity analyses with pleiotropy-robust two-sample summary-level MR approaches, including the weighted median MR^34^ and MR-Egger^35^ to compare the MR estimates between different MR models. Each of these methods provides a statistically consistent estimator of the true causal estimate under different assumptions. The intercept of the MR-Egger model represents a test for directional pleiotropy and we included this in sensitivity analyses.^35^

##### Pleiotropic pathways – inflammation and cardiometabolic risk factors

We further performed multivariable MR to adjust for potential pleiotropic pathways via cardiometabolic risk factors that are known to affect risk of both Covid-19 and CVD,^12^ including obesity (BMI),^26^ lifetime smoking index,^27^ and chronic inflammation (estimated using CRP). Multivariable MR includes all the respective genetic associations in a joint model to account for genetic confounding.^36^ While univariable MR measures the total estimate of an exposure, multivariable MR measures the direct estimate of the exposure independent of other risk factors (i.e. pleiotropy or genetic confounders) in the model.^37^ In the multivariable MR model, we selected instruments based on the primary exposure of critical Covid-19. We compared the multivariable MR model with the univariable MR model using likelihood ratio test to evaluate if accounting for the pleiotropic pathway provides a better model fit than the univariable MR model.

##### Bidirectional MR

For CVD outcomes that showed evidence of genetic correlation with critical Covid-19 in LDSC, bidirectional MR was also performed to investigate for any association of genetic liability to that CVD outcome with risk of critical Covid-19. Uncorrelated genetic variants (*r*^*2*^*<0*.*01*) associated with the CVD outcome at a *p*-value *<5* ×10^*-6*^ were selected as instruments.

MR estimates are expressed as odds ratios (OR) per unit increase in the logOR of the exposure for binary traits. All analyses were performed using the ieugwasr (version 0.1.5) and MendelianRandomization (version 0.5.0) R packages.^38^

## Results

### LD score regression

Performing LDSC, we found evidence of genetic correlation between critical Covid-19 and ischemic stroke (r_g_=0.29, FDR-*p*-value*=*4.65×10^−3^) (**Figure 1**). Critical Covid-19 was also genetically correlated with BMI (r_g_=0.21, FDR-*p*-value=6.26×10^−6^) and CRP (r_g_=0.20, FDR-*p*-value=1.35×10^−4^)). We did not observe evidence for genetic correlation between critical Covid-19 and the CVD outcomes (**Supplementary Table 2**), and therefore focused the consequent MR analysis only on ischemic stroke and its subtypes.

**Figure 1.**
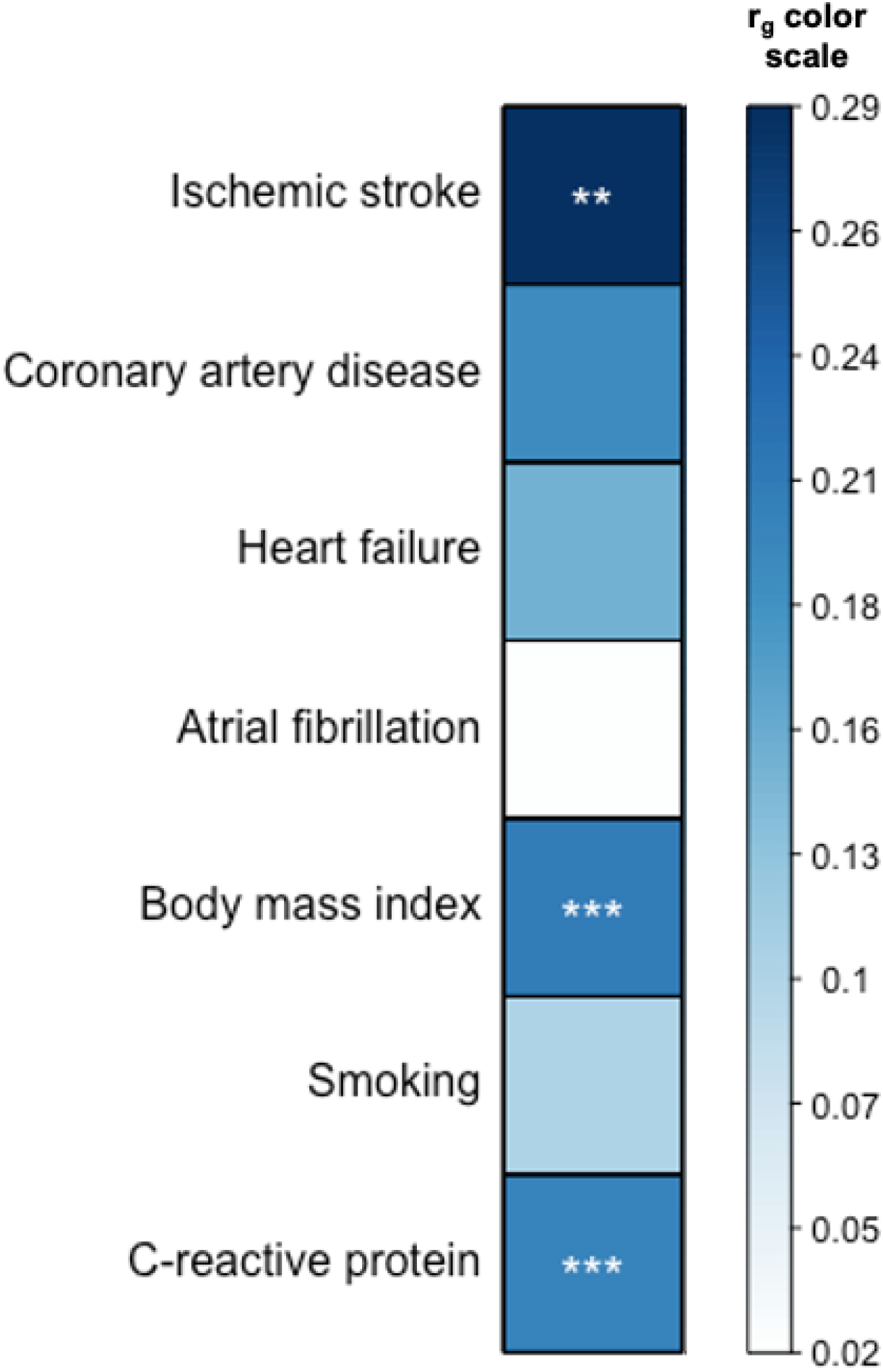
Genetic correlation between critical Covid-19 and ischemic stroke, related cardiovascular disease outcomes and risk factors for both Covid-19 and cardiovascular disease (*y*-axis), estimated by cross-trait linkage disequilibrium score regression. Asterisks indicate false discovery (FDR) corrected significant correlations (*** < 0.001, ** < 0.01, * < 0.05).

### Mendelian randomization

We selected 31 uncorrelated genetic variants as instrumental variables for liability to critical Covid-19. These are detailed in **Supplementary Table 3**, along with their associations with ischemic stroke and its subtypes. MR estimates are presented in **Figure 2**. In a univariable MR analysis, genetically proxied liability to critical Covid-19 was associated with all-cause ischemic stroke (OR 1.03, 95% CI 1.00 to 1.06, *p*-value=0.03). Restricting to ischemic stroke subtypes, there were similar MR estimates for cardioembolic stroke (OR 1.06, 95% CI 1.01 to 1.12, *p*-value=0.03), large artery stroke (OR 1.07, 95% CI 1.00 to 1.14, *p*-value=0.06) and small-vessel stroke (OR 1.05, 95% CI 1.00 to 1.11, *p*-value=0.06). For the MR estimates generated by different variants, we observed heterogeneity greater than would be expected by chance only for cardioembolic stroke (heterogeneity *p*-value=0.049), but none of the other considered ischemic stroke categories.

**Figure 2.**
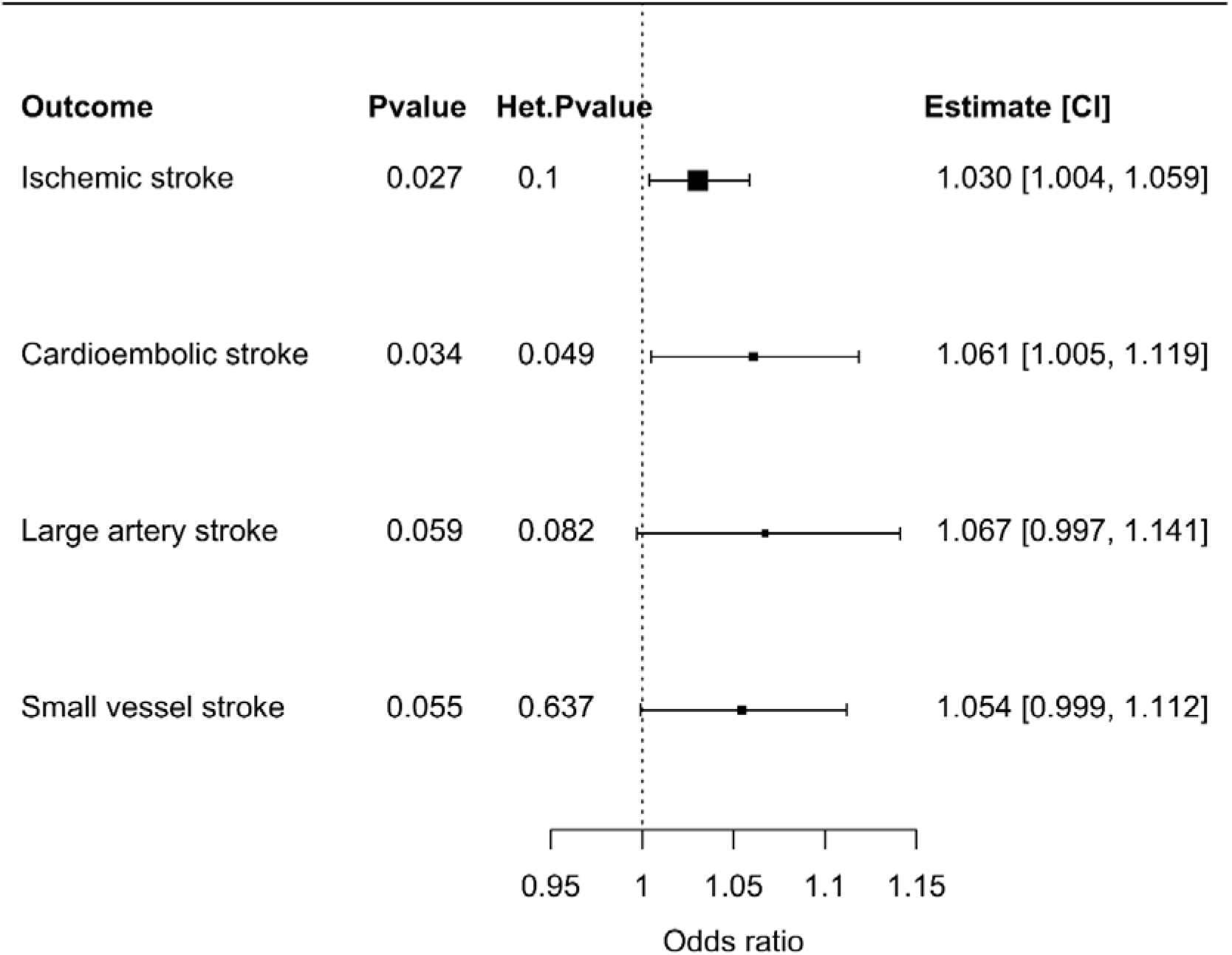
Forest plot illustrating the Mendelian randomization estimates of liability to critical Covid-19 with stroke outcomes based on inverse-variance weighted Mendelian randomization using genetic variants that were associated with critical Covid-19 at a *p*-value level of 5×10^−6^ or smaller. Mendelian randomization estimates represent the odds ratio of ischemic stroke outcomes per unit increase in the log-odds ratio of liability to critical Covid-19. Additional columns include the *p*-value of the inverse-variance weighted estimate to be different from the null (*p*-value), heterogeneity of the Mendelian randomization model measured by the Q-statistic and the respective heterogeneity p-value (Het *p*-value) as well as the Mendelian randomization estimate and its 95% confidence interval (CI). Outcomes included any ischemic stroke, cardioembolic stroke, large artery stroke, and small vessel stroke. Significant results (*p*-value < 0.05) are highlighted in bold.

### Mendelian randomization sensitivity analyses

Diagnostic scatterplots for ischemic stroke outcomes are presented in **Supplementary Figure 1**. We observed consistent MR estimates for ischemic stroke risk in sensitivity analyses based on pleiotropy-robust approaches as in the main analysis and none of the intercept estimates of MR-Egger suggested directional pleiotropy (**Supplementary Table 4**). In multivariable MR to investigate potential pleiotropy through risk factors common to both Covid-19 and CVD, there was little evidence for attenuation of the size of the estimate in any of these analyses (**Supplementary Figure 2**), which was confirmed by likelihood ratio test, (**Supplementary Table 5**).

### Sensitivity analysis based on genome-wide significant genetic variants

As an additional sensitivity analysis, we used a more stringent *p-*value threshold based on genome-wide significance to select genetic variants as instrumental variables. We identified 9 uncorrelated genetic variants that associated with critical Covid-19 at genome-wide significance (*p-*value <*5*×10^*-8*^). This MR analysis based on fewer variants generated consistent estimates to the main analysis, but with wider confidence intervals that crossed the null, reflective of lower statistical power. Results are displayed in **Supplementary Figure 3**.

### Comparison with other Covid-19 definitions

We further considered other Covid-19 definitions (**Supplementary Figure 4 and Supplementary Table 6**). Genetically predicted Covid-19 requiring hospitalization as compared to not requiring hospitalization was associated with increased risk of any ischemic stroke (OR 1.05, 95% CI 1.01 to 1.10, *p*-value=0.01) and small-vessel stroke (OR 1.22, 95% CI 1.11 to 1.34, *p*-value=5.5×10^−5^). Considering reported Covid-19 (laboratory confirmed, physician-reported or self-reported) versus controls from the general population, this was associated with increased risk of any ischemic stroke (OR 1.13, 95% CI 1.01 to 1.26, *p*-value=0.04) and large artery stroke (OR 1.46, 95% CI 1.18 to 1.81, *p*-value=4.2×10^−4^).

### Bidirectional MR

There was no strong evidence to support that genetic liability to any of the considered ischemic stroke outcomes was associated with increased risk of critical Covid-19, as illustrated in **Figure 3**.

**Figure 3.**
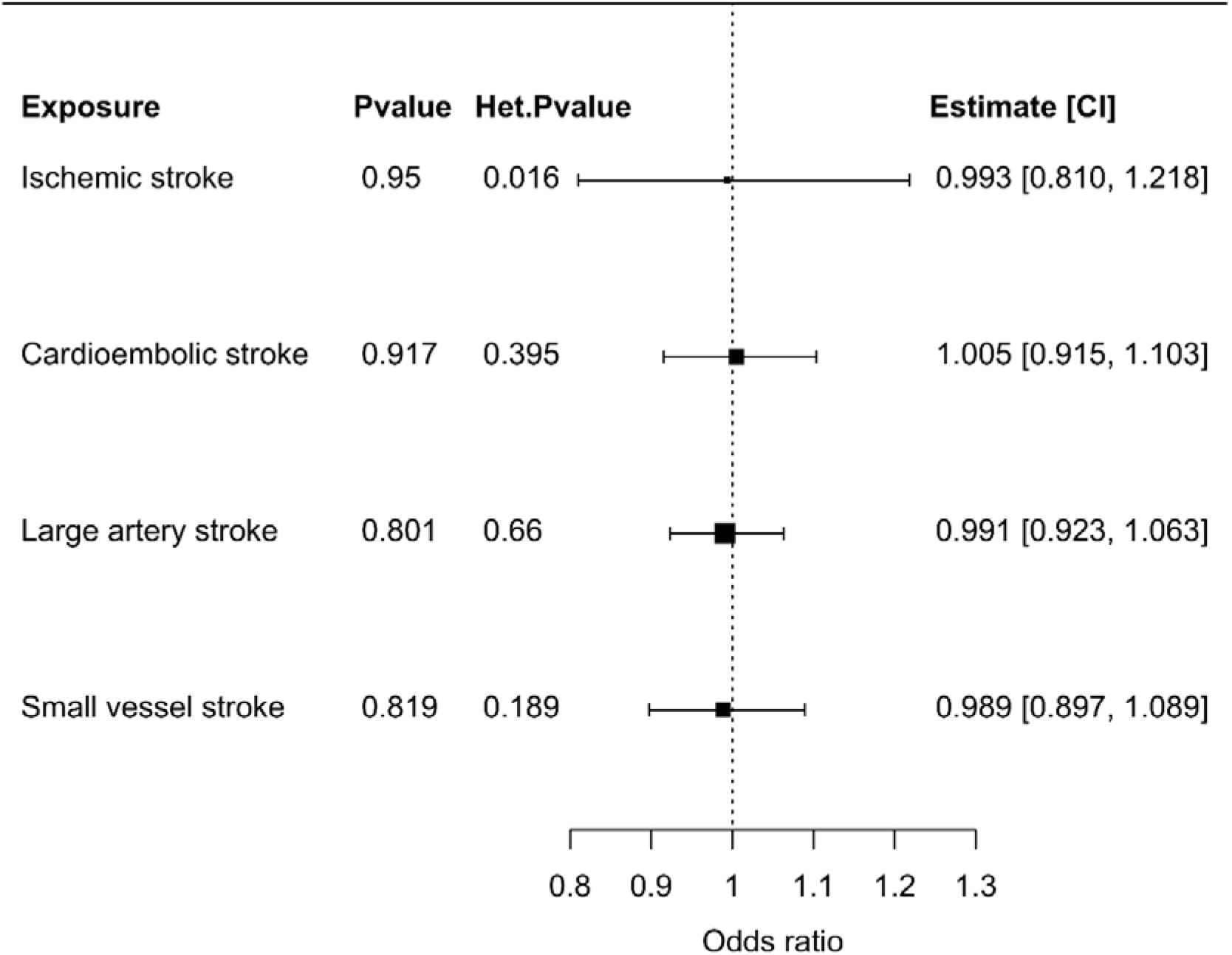
Forest plots of the bidirectional Mendelian randomization analysis illustrating the inverse-variance weighted Mendelian randomization estimate of liability to stroke phenotypes with critical Covid-19. Genetic variants which were associated with the stroke phenotypes were selected as instrumental variables at a *p*-value level of 5×10^−6^ or smaller. Mendelian randomization estimates represent the odds ratio of critical Covid-19 per unit increase in the log odds ratio of stroke phenotype. Additional columns include the *p*-value of the inverse-variance weighted Mendelian randomization estimate to be different from 1 (*p*-value), heterogeneity measured by the Q-statistic and the respective heterogeneity *p*-value (Het *p*-value), the Mendelian randomization estimate, and its 95% confidence interval (CI). Exposures included any ischemic stroke, cardioembolic stroke, large artery stroke, and small vessel stroke.

## Discussion

In this study, we used cross-trait LDSC to explore the genetic correlation of critical Covid-19 with ischemic stroke, other CVD outcomes, and risk factors common to both. We identified a genetic correlation between critical Covid-19 and ischemic stroke, and performed MR analyses that found genetic liability to critical Covid-19 to be associated with increased risk of ischemic stroke. Notably, there was no evidence to support that these associations were attributable to shared risk factors, such as obesity, smoking and chronic inflammation. Furthermore, there was no MR evidence that genetic liability to ischemic stroke increases risk of critical Covid-19.

To date, studies assessing the incidence of ischemic stroke during the Covid-19 pandemic have produced contrasting findings. On one hand, some studies demonstrate that the likelihood of stroke is seven-fold higher in people with Covid-19 than with influenza,^3^ that Covid-19 is associated with 21-fold increased odds of in-hospital stroke compared to patients without Covid-19,^6^ and that stroke is the most common neurological/neuropsychiatric complication of Covid-19.^4^ On the contrary, other studies have demonstrated a reduced rate of hospital admissions with stroke during the first wave of the pandemic compared to one year before.^9^ Two main hypotheses have been proposed as explanations for these contrasting findings. The first is that the incidence of stroke declined during the first wave of the pandemic and that Covid-19 is not mechanistically associated with stroke, and the second is that the observed reduction in stroke presentations was due to a higher proportion of people with mild strokes not reaching stroke services.^11, 39^ We have leveraged large-scale genetic data to identify that liability to critical Covid-19 is associated with increased risk of ischemic stroke. This is consistent with the host response in Covid-19 contributing to increased ischemic stroke risk. Mechanisms that increase risk of ischemic stroke in patients with Covid-19 are complex,^5, 15^ and include systemic inflammation and endotheliopathy.^15, 40–42^ Covid-19 can trigger a cytokine storm with upregulation of pro-inflammatory cytokines and chemokines such as tumor necrosis factor-α (TNF-α), interleukin-1 (IL-1) and IL-6.^15^ Endothelial inflammation can induce a microvascular and macrovascular endotheliopathy that contributes to a pro-thrombotic state.^15, 40^

While prophylactic low molecular weight heparin is used to prevent thromboembolism in patients with Covid-19, more targeted approaches to prevent strokes are yet undefined.^5, 43^ Moreover, the REMAP-CAP, ACTIV-4 and ATTACC trials have recently reported that therapeutic doses of anticoagulation do not improve clinical outcome and may increase bleeding for people with Covid-19 in the critical care setting. Previous work using an MR approach anticipated a beneficial effect of IL-6 receptor inhibition on both risk of ischemic stroke and severe Covid-19.^44, 45^ More recently, clinical trials have demonstrated that IL-6 receptor inhibition can improve outcomes in patients hospitalized with Covid-19.^46^ Targeting the deleterious host immune response through similar approaches may also help to reduce the risk of ischemic stroke and should be further evaluated.

Our findings also support the hypothesis that few patients with minor strokes reached stroke services during the first wave of the Covid-19 pandemic.^11^ This is reinforced by data that demonstrate the reduction in stroke admissions observed in some centers during the first wave of the pandemic was driven mainly by a reduction in presentations with minor stroke syndromes.^11^ People with minor stroke are at high risk of early recurrence^47^ and public health messaging should encourage people to attend stroke services if they have any symptom of stroke during the Covid-19 pandemic.

Our current study has strengths. We have made efficient use of existing large-scale data resources to address an important clinical issue in the context of the rapidly evolving global pandemic. A key strength of MR analysis is the use of randomly allocated genetic variants to help overcome environmental confounding, which is analogous to randomization of treatment allocation in clinical trials. This has helped to overcome some of the limitations of previous observational studies (either retrospective or cross-sectional) assessing the relationship between Covid-19 and ischemic stroke.^3–6^

Our work also has limitations. A series of modelling assumptions are made when using MR, in particular, that the genetic variants do not affect the considered outcomes through pathways independent of the exposure. While this can never be completely excluded, we employed methods that are robust to genetic confounding (pleiotropy) in a series of sensitivity analyses (including pleiotropy-robust MR methods and accounting for measured pleiotropy using multivariable MR) and the estimates were consistent with our main analyses. We cannot be certain that genetic associations with liability to critical Covid-19 accurately reflect the pathophysiological process that actually occurs during critical Covid-19. For example, while genetic predisposition may place an individual at increased liability to critical Covid-19, it is not possible to determine from our analyses whether that factor is involved in the pathophysiological response to Covid-19.

In conclusion, we have found genetic evidence that liability to critical Covid-19 is associated with increased risk of ischemic stroke. Our results are consistent with the host response in critical Covid-19 underlying this relationship, and support the evaluation of strategies to mitigate this.

## Data Availability

All genetic association data used in this work are publicly accessible. Statistical code related to the analyses performed in the current study is freely available from Github (https://github.com/verena-zuber/covid19_and_stroke).

## Abbreviations

BMI: Body mass index
CI: Confidence interval
Covid-19: Coronavirus disease 2019
CRP: C-reactive protein
CVD: Cardiovascular disease
FDR: False discovery rate
IVW: Inverse-variance weighted
LDSC: Linkage disequilibrium score regression
MR: Mendelian randomization
OR: Odds ratios
SARS-CoV-2: Severe acute respiratory syndrome coronavirus 2

## Acknowledgments

This work is dedicated to the memory of Maria Mion who died from Covid-19 related complications. Leonardo Bottolo is very grateful to the doctors, nurses and staff of the Ospedale di Oderzo (TV, Italy) for the high standard of care for his mother.

## References

1. Carter P, Anderson M, Mossialos E. Health system, public health, and economic implications of managing covid-19 from a cardiovascular perspective. Eur Heart J. 2020;41:2516–2518

2. Clerkin KJ, Fried JA, Raikhelkar J, Sayer G, Griffin JM, Masoumi A, et al. Covid-19 and cardiovascular disease. Circulation. 2020;141:1648–1655

3. Merkler AE, Parikh NS, Mir S, Gupta A, Kamel H, Lin E, et al. Risk of ischemic stroke in patients with coronavirus disease 2019 (covid-19) vs patients with influenza. JAMA Neurol. 2020

4. Varatharaj A, Thomas N, Ellul MA, Davies NWS, Pollak TA, Tenorio EL, et al. Neurological and neuropsychiatric complications of covid-19 in 153 patients: A uk-wide surveillance study. The Lancet Psychiatry. 2020;7:875–882

5. Qureshi AI, Abd-Allah F, Alsenani F, Aytac E, Borhani-Haghighi A, Ciccone A, et al. Management of acute ischemic stroke in patients with covid-19 infection: Report of an international panel. International Journal of Stroke. 2020;15:540–554

6. Katz JM, Libman RB, Wang JJ, Sanelli P, Filippi CG, Gribko M, et al. Cerebrovascular complications of covid-19. Stroke. 2020;51:E227–E231

7. Fuentes B, Alonso de Lecinana M, Garcia-Madrona S, Diaz-Otero F, Aguirre C, Calleja P, et al. Stroke acute management and outcomes during the covid-19 outbreak: A cohort study from the madrid stroke network. Stroke. 2021;52:552–562

8. Li Y, Li M, Wang M, Zhou Y, Chang J, Xian Y, et al. Acute cerebrovascular disease following covid-19: A single center, retrospective, observational study. Stroke Vasc Neurol. 2020;5:279–284

9. Sacco S, Ricci S, Ornello R, Eusebi P, Petraglia L, Toni D, et al. Reduced admissions for cerebrovascular events during covid-19 outbreak in italy. Stroke. 2020;51:3746–3750

10. Qureshi AI, Baskett WI, Huang W, Shyu D, Myers D, Raju M, et al. Acute ischemic stroke and covid-19: An analysis of 27 676 patients. Stroke. 2021:STROKEAHA120031786

11. Perry R, Banaras A, Werring DJ, Simister R. What has caused the fall in stroke admissions during the covid-19 pandemic? J Neurol. 2020;267:3457–3458

12. Ponsford MJ, Gkatzionis A, Walker VM, Grant AJ, Wootton RE, Moore LSP, et al. Cardiometabolic traits, sepsis, and severe covid-19: A mendelian randomization investigation. Circulation. 2020;142:1791–1793

13. Shi KB, Wood K, Shi FD, Wang XY, Liu Q. Stroke-induced immunosuppression and poststroke infection. Stroke Vasc Neurol. 2018;3:34–41

14. Didelez VSN. <mendelian randomization as an instrumental variable approach to causal inference.Pdf>.

15. Nishiga M, Wang DW, Han YL, Lewis DB, Wu JC. Covid-19 and cardiovascular disease: From basic mechanisms to clinical perspectives. Nat Rev Cardiol. 2020;17:543–558

16. Initiative C-HG. The covid-19 host genetics initiative, a global initiative to elucidate the role of host genetic factors in susceptibility and severity of the sars-cov-2 virus pandemic. Eur J Hum Genet. 2020;28:715–718

17. <stroke and subtypes.Pdf>.

18. Malik R, Chauhan G, Traylor M, Sargurupremraj M, Okada Y, Mishra A, et al. Multiancestry genome-wide association study of 520,000 subjects identifies 32 loci associated with stroke and stroke subtypes. Nat Genet. 2018;50:524–537

19. Nikpay M, Goel A, Won HH, Hall LM, Willenborg C, Kanoni S, et al. A comprehensive 1,000 genomes-based genome-wide association meta-analysis of coronary artery disease. Nat Genet. 2015;47:1121–1130

20. Shah S, Henry A, Roselli C, Lin HH, Sveinbjornsson G, Fatemifar G, et al. Genome-wide association and mendelian randomisation analysis provide insights into the pathogenesis of heart failure. Nat Commun. 2020;11

21. Roselli C, Chaffin MD, Weng LC, Aeschbacher S, Ahlberg G, Albert CM, et al. Multi-ethnic genome-wide association study for atrial fibrillation. Nat Genet. 2018;50:1225-+

22. Lindsberg PJ, Grau AJ. Inflammation and infections as risk factors for ischemic stroke. Stroke. 2003;34:2518–2532

23. Toyoda K, Ninomiya T. Stroke and cerebrovascular diseases in patients with chronic kidney disease. Lancet Neurol. 2014;13:823–833

24. Kernan WN, Inzucchi SE, Sawan C, Macko RF, Furie KL. Obesity a stubbornly obvious target for stroke prevention. Stroke. 2013;44:278–286

25. Wolf PA, Dagostino RB, Kannel WB, Bonita R, Belanger AJ. Cigarette-smoking as a risk factor for stroke - the framingham-study. Jama-J Am Med Assoc. 1988;259:1025–1029

26. Pulit SL, Stoneman C, Morris AP, Wood AR, Glastonbury CA, Tyrrell J, et al. Meta-analysis of genome-wide association studies for body fat distribution in 694 649 individuals of european ancestry. Hum Mol Genet. 2019;28:166–174

27. Wootton RE, Richmond RC, Stuijfzand BG, Lawn RB, Sallis HM, Taylor GMJ, et al. Evidence for causal effects of lifetime smoking on risk for depression and schizophrenia: A mendelian randomisation study. Psychol Med. 2020;50:2435–2443

28. Neale lab. Accessed 2020 june 16. Rapid gwas of thousands of phenotypes in the uk biobank 2020. http://www.Nealelab.Is/uk-biobank/ukbround2announcement.

29. Wuttke M, Li Y, Li M, Sieber KB, Feitosa MF, Gorski M, et al. A catalog of genetic loci associated with kidney function from analyses of a million individuals. Nat Genet. 2019;51:957-+

30. Bulik-Sullivan BK, Loh PR, Finucane HK, Ripke S, Yang J, Patterson N, et al. Ld score regression distinguishes confounding from polygenicity in genome-wide association studies. Nat Genet. 2015;47:291-+

31. Benjamini Y, Hochberg Y. Controlling the false discovery rate - a practical and powerful approach to multiple testing. J R Stat Soc B. 1995;57:289–300

32. Pierce BL, Burgess S. Efficient design for mendelian randomization studies: Subsample and 2-sample instrumental variable estimators. Am J Epidemiol. 2013;178:1177–1184

33. Greco MF, Minelli C, Sheehan NA, Thompson JR. Detecting pleiotropy in mendelian randomisation studies with summary data and a continuous outcome. Stat Med. 2015;34:2926–2940

34. Bowden J, Davey Smith G, Haycock PC, Burgess S. Consistent estimation in mendelian randomization with some invalid instruments using a weighted median estimator. Genet Epidemiol. 2016;40:304–314

35. Bowden J, Davey Smith G, Burgess S. Mendelian randomization with invalid instruments: Effect estimation and bias detection through egger regression. Int J Epidemiol. 2015;44:512–525

36. Burgess S, Thompson SG. Multivariable mendelian randomization: The use of pleiotropic genetic variants to estimate causal effects. Am J Epidemiol. 2015;181:251–260

37. Burgess S, Thompson DJ, Rees JMB, Day FR, Perry JR, Ong KK. Dissecting causal pathways using mendelian randomization with summarized genetic data: Application to age at menarche and risk of breast cancer. Genetics. 2017;207:481–487

38. Yavorska OO, Burgess S. Mendelianrandomization: An r package for performing mendelian randomization analyses using summarized data. Int J Epidemiol. 2017;46:1734–1739

39. Aguiar de Sousa D, Sandset EC, Elkind MSV. The curious case of the missing strokes during the covid-19 pandemic. Stroke. 2020;51:1921–1923

40. Varga Z, Flammer AJ, Steiger P, Haberecker M, Andermatt R, Zinkernagel AS, et al. Endothelial cell infection and endotheliitis in covid-19. Lancet. 2020;395:1417–1418

41. Lee SHG, Fralick M, Sholzberg M. Coagulopathy associated with covid-19. Can Med Assoc J. 2020;192:E583–E583

42. Fauvel C, Weizman O, Trimaille A, Mika D, Pommier T, Pace N, et al. Pulmonary embolismin covid-19 patients: A french multicentre cohort study. European Heart Journal. 2020;41:3058–3068

43. Robinson RG, Bolduc PL, Price TR. Two-year longitudinal study of poststroke mood disorders: Diagnosis and outcome at one and two years. Stroke. 1987;18:837–843

44. Georgakis MK, Malik R, Gill D, Franceschini N, Sudlow CLM, Dichgans M, et al. Interleukin-6 signaling effects on ischemic stroke and other cardiovascular outcomes a mendelian randomization study. Circ-Genom Precis Me. 2020;13:168–171

45. Larsson SC, Burgess S, Gill DC. Genetically proxied inhibition of interleukin-6 signaling: Opposing associations with susceptibility to covid-19 and pneumonia. 2020

46. Salama C, Han J, Yau L, Reiss WG, Kramer B, Neidhart JD, et al. Tocilizumab in patients hospitalized with covid-19 pneumonia. N Engl J Med. 2021;384:20–30

47. Coull AJ, Lovett JK, Rothwell PM, Oxford Vascular S. Population based study of early risk of stroke after transient ischaemic attack or minor stroke: Implications for public education and organisation of services. BMJ. 2004;328:326

