## Supplement for "Leveraging genetic data to elucidate the relationship between Covid-19 and ischemic stroke"

Short title: Covid-19 and ischemic stroke

Verena Zuber*, Alan Cameron*, Pavlos Myserlis, Leonardo Bottolo, Israel Fernandez, Stephen Burgess, Chris Anderson, Jesse Dawson, Dipender Gill

*These authors contributed equally and are joint first

**Supplementary Table 1**: Overview of the publicly available summary-level data of genetic associations used for the analysis.

| Phenotype | Description | Sample Size | Cases | Controls | Population | Pubmed | Author |
| --- | --- | --- | --- | --- | --- | --- | --- |
| Covid-19 | Criticall Covid *vs.* population |  | 5,101 | 1,383,241 | EUR |  | Covid19-HG |
|  | Hospitalized Covid *vs.* not hospitalized Covid |  | 4,829 | 11,816 |  |  |  |
|  | Hospitalized Covid *vs.* population |  | 9,986 | 1,877,672 |  |  |  |
|  | Covid vs population |  | 38,984 | 1,644,784 |  |  |  |
| Stroke | Any ischemic stroke |  | 34,217 | 406,111 | EUR | 29531354 | Malik et al. (2018) |
|  | Large artery stroke |  | 4,373 | 406,111 |  |  |  |
|  | Cardioembolic stroke |  | 7,193 | 406,111 |  |  |  |
|  | Small vessel stroke |  | 5,386 | 406,111 |  |  |  |
| Cardiovascular disease outcomes | Coronary artery disease |  | 60,801 | 123,504 | EUR, SEA | 26343387 | Nikpay et al. (2015) |
|  | Heart failure |  | 47,309 | 930,014 | EUR | 31919418 | Shah et al. (2020) |
|  | Atrial fibrillation |  | 65,446 | 522,744 | TRANS | 29892015 | Roselli et al. (2018) |
| Obesity | Body mass index | 694,649 |  | | EUR | 30239722 | Pullit et al. (2019) |
| Smoking | Lifetime smoking index | 462,690 |  |  | EUR | 30239722 | Wootton et al. (2020) |
| Inflammation | C-reactive protein | 361,194 |  |  | EUR |  | Neale lab |

**Supplementary Table 2**. Cross-trait linkage disequilibrium score regression (LDSC) analysis results of critical Covid-19 with ischemic stroke, cardiovascular disease (CVD) outcomes, and risk factors related to both Covid-19 and CVD. r_g_ represents the genetic correlation between critical Covid-19 and each phenotype. *p*-values are corrected with the Benjamini and Hochberg false discovery rate (FDR).

|  | **r_g_** | **FDR-*p* value** |
| --- | --- | --- |
| **Ischemic stroke** | 0.2922 | 4.65E-03 |
| **Coronary artery disease** | 0.1914 | 7.60E-02 |
| **Heart failure** | 0.1491 | 1.12E-01 |
| **Atrial fibrillation** | 0.0198 | 7.17E-01 |
| **Body mass index** | 0.2088 | 6.26E-06 |
| **Smoking** | 0.1029 | 5.91E-02 |
| **C-reactive protein** | 0.2006 | 1.35E-04 |

**Supplementary Table 3**: Overview of the genetic variants used as instrumental variables for liability to critical Covid-19 based on the Covid-19 host genetics initiative. We selected 31 uncorrelated (clumped at correlation threshold *r^2^ < 0.01*) genetic variants as instrumental variables for laibility to critical Covid-19 that were associated at a *p*-value level of *5 × 10^-6^* or smaller*.* The table additionally includes summary-level data (beta coefficients of genetic association, their standard error and corresponding *p-*value) for liability to critical Covid-19 as exposure and any ischemic stroke (AIS) and its subtypes cardioembolic stroke (CES), large artery stroke (LAS), and small vessel stroke (SVS) as outcomes. Alt: alternative allele; Chr: chromosome; Pos: position; Ref: reference allele; SE: standard error.

|  | | | | | **Critical Covid-19** | | | **AIS** | | | **CES** | | | **LAS** | | | **SVS** | | |
| --- | --- | --- | --- | --- | --- | --- | --- | --- | --- | --- | --- | --- | --- | --- | --- | --- | --- | --- | --- |
| **Variant** | **Chr** | **Pos** | **Ref** | **Alt** | **Beta** | **SE** | **P** | **Beta** | **SE** | **P** | **Beta** | **SE** | **P** | **Beta** | **SE** | **P** | **Beta** | **SE** | **P** |
| rs10087754 | 8 | 121819908 | T | A | -1.3E-01 | 2.7E-02 | 7.1E-07 | -1.2E-02 | 1.0E-02 | 2.4E-01 | 1.0E-04 | 1.9E-02 | 9.9E-01 | -4.2E-02 | 2.5E-02 | 9.6E-02 | -9.6E-03 | 2.3E-02 | 6.8E-01 |
| rs11085727 | 19 | 10355447 | C | T | 1.7E-01 | 2.9E-02 | 3.7E-09 | -2.2E-02 | 1.1E-02 | 5.2E-02 | -5.5E-02 | 2.2E-02 | 1.1E-02 | -5.0E-04 | 2.8E-02 | 9.8E-01 | 3.5E-02 | 2.5E-02 | 1.7E-01 |
| rs111508230 | 1 | 155181061 | C | T | -2.1E-01 | 4.4E-02 | 2.6E-06 | -8.1E-03 | 1.6E-02 | 6.1E-01 | -7.1E-02 | 3.1E-02 | 2.4E-02 | 9.3E-02 | 3.9E-02 | 1.7E-02 | 2.9E-02 | 4.1E-02 | 4.8E-01 |
| rs114969787 | 5 | 65770656 | C | T | 3.1E-01 | 6.6E-02 | 3.6E-06 | 4.1E-02 | 3.1E-02 | 1.9E-01 | -4.7E-02 | 6.4E-02 | 4.7E-01 | 1.2E-01 | 7.7E-02 | 1.4E-01 | 4.4E-02 | 7.2E-02 | 5.5E-01 |
| rs11658357 | 17 | 36097317 | A | T | -2.0E-01 | 4.4E-02 | 4.9E-06 | 9.7E-03 | 1.3E-02 | 4.4E-01 | -1.8E-02 | 2.4E-02 | 4.5E-01 | -1.5E-02 | 3.1E-02 | 6.1E-01 | 4.4E-02 | 2.9E-02 | 1.3E-01 |
| rs117232645 | 13 | 74553195 | G | A | -3.3E-01 | 7.0E-02 | 2.5E-06 | -3.1E-02 | 3.0E-02 | 3.0E-01 | 1.0E-02 | 5.9E-02 | 8.6E-01 | 4.4E-02 | 7.3E-02 | 5.4E-01 | 3.8E-03 | 6.8E-02 | 9.6E-01 |
| rs13050728 | 21 | 33242905 | T | C | -2.0E-01 | 2.9E-02 | 2.4E-12 | -4.4E-03 | 1.1E-02 | 6.8E-01 | -2.9E-02 | 2.0E-02 | 1.5E-01 | -2.7E-02 | 2.6E-02 | 3.1E-01 | 2.7E-02 | 2.5E-02 | 2.8E-01 |
| rs13080258 | 3 | 69672908 | A | C | -1.4E-01 | 3.0E-02 | 3.6E-06 | 2.8E-03 | 1.2E-02 | 8.1E-01 | 6.8E-03 | 2.3E-02 | 7.6E-01 | -1.6E-02 | 2.9E-02 | 5.9E-01 | -1.8E-02 | 2.7E-02 | 5.1E-01 |
| rs13274496 | 8 | 22583385 | G | A | -2.0E-01 | 4.2E-02 | 1.8E-06 | 7.0E-04 | 1.3E-02 | 9.6E-01 | 1.7E-02 | 2.4E-02 | 4.8E-01 | -1.9E-02 | 3.2E-02 | 5.5E-01 | -3.2E-02 | 2.9E-02 | 2.6E-01 |
| rs143334143 | 6 | 31153649 | G | A | 2.9E-01 | 4.3E-02 | 6.0E-12 | 9.0E-03 | 1.9E-02 | 6.4E-01 | 3.6E-02 | 3.6E-02 | 3.1E-01 | -1.0E-02 | 4.8E-02 | 8.3E-01 | 3.6E-02 | 4.6E-02 | 4.3E-01 |
| rs1974792 | 19 | 50353078 | A | G | -1.4E-01 | 2.7E-02 | 5.1E-07 | -2.0E-02 | 1.0E-02 | 4.7E-02 | -9.6E-03 | 2.0E-02 | 6.3E-01 | -6.3E-02 | 2.5E-02 | 1.3E-02 | 3.2E-02 | 2.4E-02 | 1.8E-01 |
| rs2109069 | 19 | 4719431 | G | A | 2.6E-01 | 2.8E-02 | 6.1E-20 | -7.1E-03 | 1.1E-02 | 5.2E-01 | 2.0E-03 | 2.2E-02 | 9.3E-01 | -6.2E-03 | 2.7E-02 | 8.2E-01 | -1.1E-02 | 2.6E-02 | 6.8E-01 |
| rs2237698 | 7 | 107967457 | C | T | 2.4E-01 | 4.0E-02 | 2.4E-09 | -2.7E-02 | 2.1E-02 | 1.9E-01 | -2.7E-02 | 3.9E-02 | 4.8E-01 | 4.8E-02 | 5.1E-02 | 3.5E-01 | -2.3E-02 | 4.7E-02 | 6.3E-01 |
| rs2597569 | 11 | 97922951 | T | C | -1.8E-01 | 3.4E-02 | 9.3E-08 | -7.0E-03 | 1.0E-02 | 4.9E-01 | -1.5E-02 | 1.9E-02 | 4.5E-01 | -3.3E-02 | 2.5E-02 | 2.0E-01 | -3.5E-02 | 2.4E-02 | 1.4E-01 |
| rs2733839 | 12 | 10393411 | T | C | 2.7E-01 | 5.8E-02 | 3.5E-06 | -3.9E-03 | 2.7E-02 | 8.9E-01 | 4.3E-02 | 5.3E-02 | 4.2E-01 | -4.1E-02 | 7.2E-02 | 5.7E-01 | 3.0E-02 | 6.4E-02 | 6.4E-01 |
| rs340850 | 1 | 213941523 | T | G | -2.8E-01 | 6.0E-02 | 4.3E-06 | 2.0E-02 | 2.4E-02 | 4.0E-01 | 5.6E-02 | 4.8E-02 | 2.5E-01 | -9.7E-02 | 5.7E-02 | 9.0E-02 | -5.3E-02 | 5.5E-02 | 3.3E-01 |
| rs35081325 | 3 | 45848429 | A | T | 6.3E-01 | 4.5E-02 | 5.8E-45 | 3.0E-02 | 2.0E-02 | 1.3E-01 | 4.3E-02 | 3.8E-02 | 2.6E-01 | 5.4E-02 | 5.1E-02 | 2.8E-01 | 2.7E-02 | 4.9E-02 | 5.9E-01 |
| rs36932 | 7 | 123877938 | G | A | -1.6E-01 | 3.4E-02 | 4.0E-06 | -6.2E-03 | 1.4E-02 | 6.5E-01 | -2.1E-02 | 2.6E-02 | 4.3E-01 | -3.2E-02 | 3.3E-02 | 3.3E-01 | -4.8E-02 | 3.1E-02 | 1.2E-01 |
| rs4076440 | 1 | 9630418 | A | G | 2.1E-01 | 4.3E-02 | 9.4E-07 | -1.5E-02 | 2.1E-02 | 4.7E-01 | 4.2E-03 | 4.1E-02 | 9.2E-01 | -5.8E-02 | 5.3E-02 | 2.7E-01 | 5.1E-03 | 4.8E-02 | 9.2E-01 |
| rs5767981 | 22 | 47769327 | A | G | -1.6E-01 | 3.4E-02 | 5.0E-06 | -2.0E-02 | 1.1E-02 | 5.7E-02 | -7.9E-03 | 2.1E-02 | 7.0E-01 | -3.0E-03 | 2.7E-02 | 9.1E-01 | -2.6E-02 | 2.5E-02 | 2.9E-01 |
| rs622568 | 7 | 54580201 | A | C | 2.3E-01 | 3.7E-02 | 1.0E-09 | 3.5E-03 | 1.4E-02 | 8.0E-01 | 5.2E-02 | 2.6E-02 | 4.4E-02 | -5.7E-02 | 3.4E-02 | 9.1E-02 | -1.7E-02 | 3.1E-02 | 5.8E-01 |
| rs633862 | 9 | 133279871 | T | C | -1.7E-01 | 3.4E-02 | 1.1E-06 | -2.6E-02 | 1.0E-02 | 1.2E-02 | -5.5E-02 | 1.9E-02 | 3.8E-03 | -5.1E-02 | 2.4E-02 | 3.7E-02 | -1.0E-04 | 2.3E-02 | 1.0E+00 |
| rs6478109 | 9 | 114806486 | A | G | 1.5E-01 | 2.8E-02 | 2.4E-07 | 1.2E-02 | 1.1E-02 | 2.5E-01 | 2.2E-02 | 2.1E-02 | 2.8E-01 | 6.0E-02 | 2.7E-02 | 2.7E-02 | 4.4E-02 | 2.5E-02 | 7.4E-02 |
| rs6712600 | 2 | 125728622 | G | A | -1.7E-01 | 3.4E-02 | 3.3E-07 | 6.9E-03 | 1.3E-02 | 5.8E-01 | 5.0E-02 | 2.4E-02 | 3.7E-02 | -9.4E-03 | 3.1E-02 | 7.6E-01 | -3.1E-02 | 2.9E-02 | 2.8E-01 |
| rs7135260 | 12 | 112943944 | T | C | 1.9E-01 | 2.8E-02 | 6.1E-12 | 2.6E-02 | 1.1E-02 | 1.5E-02 | 0.0E+00 | 2.1E-02 | 1.0E+00 | 4.0E-02 | 2.7E-02 | 1.4E-01 | 4.1E-02 | 2.5E-02 | 1.0E-01 |
| rs77406469 | 7 | 150758864 | C | T | -3.4E-01 | 6.9E-02 | 6.5E-07 | 1.8E-02 | 2.7E-02 | 5.1E-01 | -4.3E-02 | 6.0E-02 | 4.7E-01 | 5.4E-02 | 7.6E-02 | 4.8E-01 | 6.6E-02 | 6.8E-02 | 3.3E-01 |
| rs77534576 | 17 | 49863303 | C | T | 4.6E-01 | 7.5E-02 | 8.5E-10 | 2.8E-02 | 3.1E-02 | 3.7E-01 | 5.4E-02 | 6.6E-02 | 4.1E-01 | 7.3E-02 | 8.2E-02 | 3.7E-01 | 1.3E-01 | 7.6E-02 | 7.6E-02 |
| rs79833209 | 5 | 163300447 | C | T | 4.4E-01 | 9.2E-02 | 2.2E-06 | 1.0E-01 | 3.6E-02 | 3.9E-03 | 8.1E-02 | 7.9E-02 | 3.0E-01 | -8.4E-03 | 9.9E-02 | 9.3E-01 | -2.0E-02 | 9.0E-02 | 8.2E-01 |
| rs9287218 | 1 | 237113798 | A | C | -3.2E-01 | 6.2E-02 | 3.9E-07 | 2.8E-03 | 2.4E-02 | 9.1E-01 | 2.3E-02 | 4.7E-02 | 6.3E-01 | 1.2E-02 | 6.2E-02 | 8.5E-01 | -4.9E-02 | 5.8E-02 | 4.0E-01 |
| rs9577175 | 13 | 112889041 | C | T | 2.0E-01 | 4.1E-02 | 7.9E-07 | -4.0E-03 | 1.2E-02 | 7.3E-01 | 3.8E-03 | 2.3E-02 | 8.7E-01 | -2.5E-02 | 3.0E-02 | 4.0E-01 | 1.2E-02 | 2.8E-02 | 6.7E-01 |
| rs9871880 | 3 | 197399535 | C | T | -2.5E-01 | 5.0E-02 | 4.1E-07 | -2.7E-03 | 1.8E-02 | 8.8E-01 | -1.0E-01 | 3.6E-02 | 4.1E-03 | 5.4E-02 | 4.4E-02 | 2.2E-01 | 1.0E-02 | 4.2E-02 | 8.1E-01 |

**Supplementary Table 4**: Sensitivity analysis for the Mendelian randomization analysis of liability to critical Covid-19 on ischemic stroke outcomes including the inverse-variance weighted (IVW) Mendelian randomization and pleiotropy-robust Mendelian randomization approaches (simple, weighted median and MR-Egger). Mendelian randomization estimates represent the odds ratio for ischemic stroke outcomes per unit increase in the log-odds ratio of liability to critical Covid-19. In addition to the Mendelian randomization estimates, we included their 95% confidence interval (CI) and corresponding *p-*value. The intercept of the MR-Egger method was used to test for directional pleiotropy. Instrument selection was based on genetic variants that were associated with liability to critical Covid-19 with a *p*-value equal to or smaller than *5 × 10^-6^*. Main outcome was any ischemic stroke, and we further included the subtypes cardioembolic stroke, large artery stroke, and small vessel stroke.

| **Outcome** | **Method** | **Estimate** | **95% CI Lower** | **95% CI Upper** | ***p*-value** |
| --- | --- | --- | --- | --- | --- |
| **Any ischemic stroke** | IVW | 1.031 | 1.004 | 1.058 | 0.027 |
|  | Simple median | 1.022 | 0.986 | 1.059 | 0.227 |
|  | Weighted median | 1.031 | 0.995 | 1.068 | 0.088 |
|  | MR-Egger | 1.022 | 0.947 | 1.103 | 0.573 |
|  | MR-Egger intercept | 0.002 | -0.014 | 0.018 | 0.820 |
| **Cardio- embolic stroke** | IVW | 1.060 | 1.005 | 1.119 | 0.034 |
|  | Simple median | 1.072 | 1.001 | 1.147 | 0.047 |
|  | Weighted median | 1.072 | 1.001 | 1.147 | 0.046 |
|  | MR-Egger | 1.092 | 0.935 | 1.276 | 0.266 |
|  | MR-Egger intercept | -0.007 | -0.040 | 0.026 | 0.689 |
| **Large artery stroke** | IVW | 1.067 | 0.997 | 1.141 | 0.059 |
|  | Simple median | 1.079 | 0.988 | 1.180 | 0.091 |
|  | Weighted median | 1.087 | 0.994 | 1.188 | 0.066 |
|  | MR-Egger | 0.919 | 0.762 | 1.108 | 0.374 |
|  | MR-Egger intercept | 0.034 | -0.006 | 0.073 | 0.095 |
| **Small vessel stroke** | IVW | 1.054 | 0.999 | 1.112 | 0.055 |
|  | Simple median | 1.075 | 0.990 | 1.167 | 0.087 |
|  | Weighted median | 1.051 | 0.968 | 1.140 | 0.237 |
|  | MR-Egger | 1.012 | 0.867 | 1.182 | 0.879 |
|  | MR-Egger intercept | 0.009 | -0.024 | 0.042 | 0.585 |

**Supplementary Table 5**: Likelihood ratio test to compare the model fit of the multivariable Mendelian randomization model considering risk for critical Covid-19 as exposure for ischemic stroke outcomes accounting for potential pleiotropic pathways (including life-time smoking index, body mass index, and c-reactive protein) with the univariable Mendelian randomization model. Model fit is evaluated using residual sum of squares for the univariable Mendelian randomization model (RSS 1) with the residual sum of squares for the multivariable Mendelian randomization model (RSS 2). There was one degree of freedom difference between the multivariable and the univariable Mendelian randomization model because there is one additional parameter to estimate in the multivariable Mendelian randomization model. The *F*-statistic quantifies the reduction in residual sum of squares by adding the pleiotropic risk factor to the Mendelian randomization model. The respective *p*-value tests if the multivariable Mendelian randomization model including the pleiotropic pathways provides a significantly better model fit of the genetic association estimates with the stroke outcome than the univariable Mendelian randomization model. Instrument selection was based on genetic variants that were associated with liability to critical Covid-19 with a *p*-value equal to or smaller than *5 × 10^-6^*. Main outcome was any ischemic stroke, and we further included the subtypes cardioembolic stroke, large artery stroke, and small vessel stroke.

| **Outcome** | **Pleiotropic pathway** | **RSS 1** | **RSS 2** | ***F*-statistic** | ***p*-value** |
| --- | --- | --- | --- | --- | --- |
| **Any ischemic stroke** | **Smoking** | 40.230 | 39.436 | 0.584 | 0.451 |
|  | **Body mass index** | 40.230 | 36.919 | 2.601 | 0.118 |
|  | **C-reactive protein** | 37.457 | 34.841 | 2.103 | 0.158 |
| **Cardio- embolic stroke** | **Smoking** | 43.899 | 42.439 | 0.998 | 0.326 |
|  | **Body mass index** | 43.899 | 43.733 | 0.110 | 0.743 |
|  | **C-reactive protein** | 42.765 | 37.544 | 3.894 | 0.058 |
| **Large artery stroke** | **Smoking** | 41.290 | 40.612 | 0.483 | 0.492 |
|  | **Body mass index** | 41.290 | 41.290 | 0.000 | 0.997 |
|  | **C-reactive protein** | 40.872 | 39.648 | 0.865 | 0.360 |
| **Small vessel stroke** | **Smoking** | 26.742 | 26.150 | 0.656 | 0.425 |
|  | **Body mass index** | 26.742 | 23.993 | 3.322 | 0.079 |
|  | **C-reactive protein** | 26.177 | 26.176 | 0.001 | 0.978 |

**Supplementary Table 6**: Mendelian randomization estimates from the inverse-variance weighted Mendelian randomization analysis considering different Covid-19 phenotypes as exposure for ischemic stroke subtypes. Covid-19 phenotypes were based on the definitions by the Covid-19 host genetics initiative. Mendelian randomization estimates represent the odds ratio for ischemic stroke outcomes per unit increase in the log-odds ratio of liability to the respective Covid-19 definition. In addition to the Mendelian randomization estimates, we included their 95% confidence interval (CI) and corresponding *p-*value. Instrument selection was based on genetic variants that were associated with the respective Covid-19 definition with a *p*-value equal to or smaller than *5 × 10^-6^*. Moreover, we displayed heterogeneity measured by the Q-statistic and the corresponding heterogeneity *p-*value (Het. *p*-value). Main outcome was any ischemic stroke, and we further included the subtypes cardioembolic stroke, large artery stroke, and small vessel stroke.

| **Exposure** | **Outcome** | **Estimate** | **95% CI Lower** | **95% CI Upper** | ***p*-value** | **Q-statistic** | **Het.**  ***p*-value** |
| --- | --- | --- | --- | --- | --- | --- | --- |
| **Hospitalized for Covid-19 *versus* controls with laboratory-confirmed Covid-19** | **Any ischemic stroke** | 1.054 | 1.012 | 1.099 | 0.011 | 5.311 | 0.915 |
|  | **Cardioembolic stroke** | 1.044 | 0.963 | 1.133 | 0.294 | 5.062 | 0.928 |
|  | **Large artery stroke** | 1.061 | 0.957 | 1.177 | 0.258 | 5.080 | 0.927 |
|  | **Small vessel stroke** | 1.219 | 1.107 | 1.342 | 5.5×10^-5^ | 7.964 | 0.717 |
| **Hospitalized for Covid-19 *versus* population controls** | **Any ischemic stroke** | 1.026 | 0.981 | 1.073 | 0.268 | 46.406 | 0.021 |
|  | **Cardioembolic stroke** | 1.090 | 0.991 | 1.198 | 0.078 | 56.523 | 0.002 |
|  | **Large artery stroke** | 1.081 | 0.978 | 1.194 | 0.128 | 37.369 | 0.137 |
|  | **Small vessel stroke** | 0.991 | 0.912 | 1.078 | 0.841 | 30.302 | 0.399 |
| **Reported Covid-19 *versus* population controls** | **Any ischemic stroke** | 1.126 | 1.005 | 1.262 | 0.041 | 59.811 | 1.1×10^-4^ |
|  | **Cardioembolic stroke** | 1.158 | 0.960 | 1.396 | 0.125 | 44.238 | 0.010 |
|  | **Large artery stroke** | 1.464 | 1.184 | 1.811 | 4.2×10^-4^ | 33.982 | 0.108 |
|  | **Small vessel stroke** | 1.043 | 0.879 | 1.237 | 0.629 | 25.093 | 0.457 |

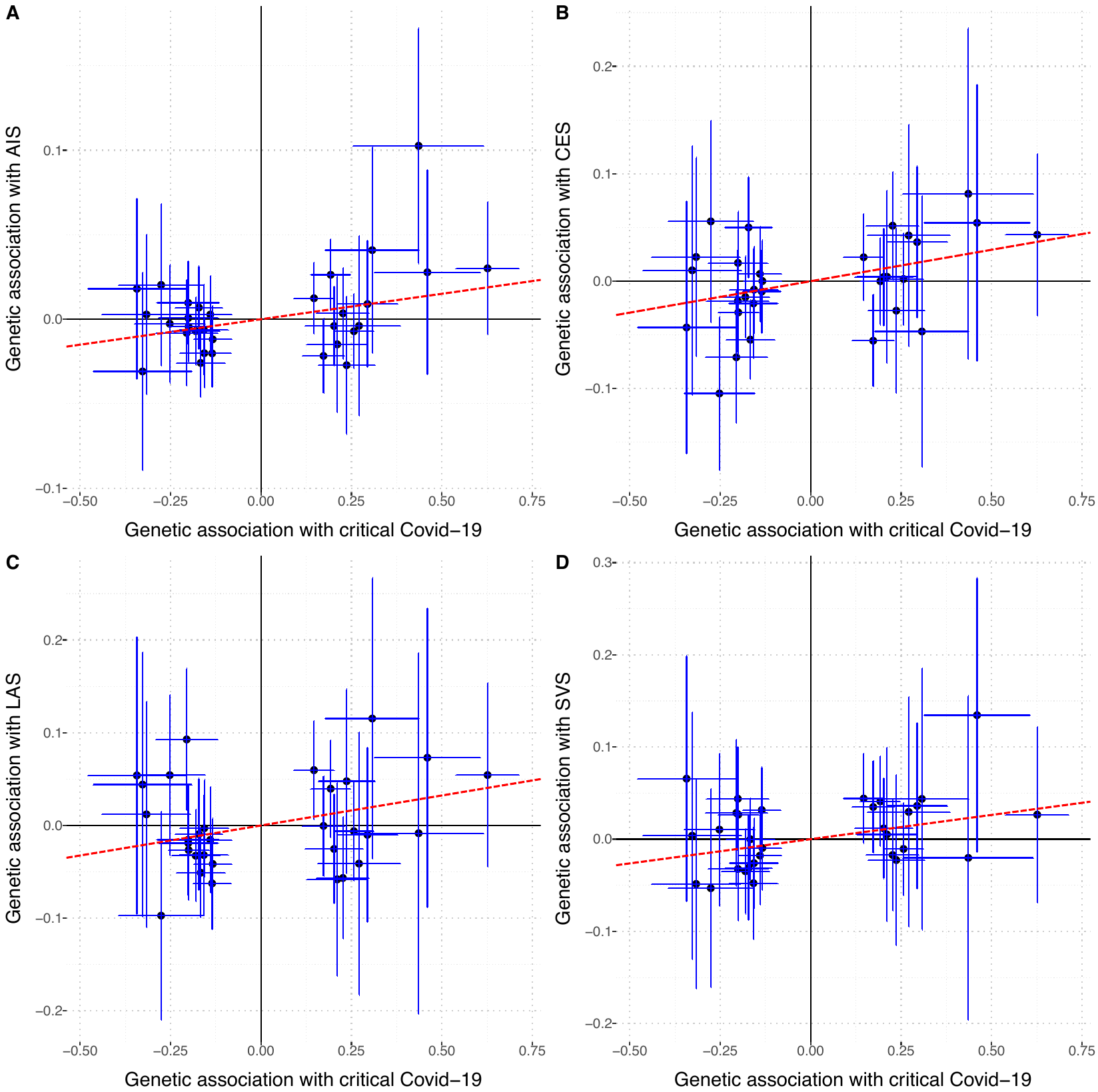

**Supplementary Figure 1**: Scatterplots for diagnostics plot the genetic association of the 31 genetic variants used as instrumental variables with the exposure (liability to critical Covid-19) on the *x*-axis against genetic associations with the outcome (ischemic stroke phenotypes) on the *y*-axis. Error bars indicate the standard error of the genetic association. The inverse-variance weighted Mendelian randomization estimate is represented by the red dashed line through the origin. Each panel shows main outcome: Panel **A**) any ischemic stroke (**AIS**), **B**) cardioembolic stroke (**CES**), **C**) large artery stroke (**LAS**), and **D**) small vessel stroke (**SVS**), respectively.

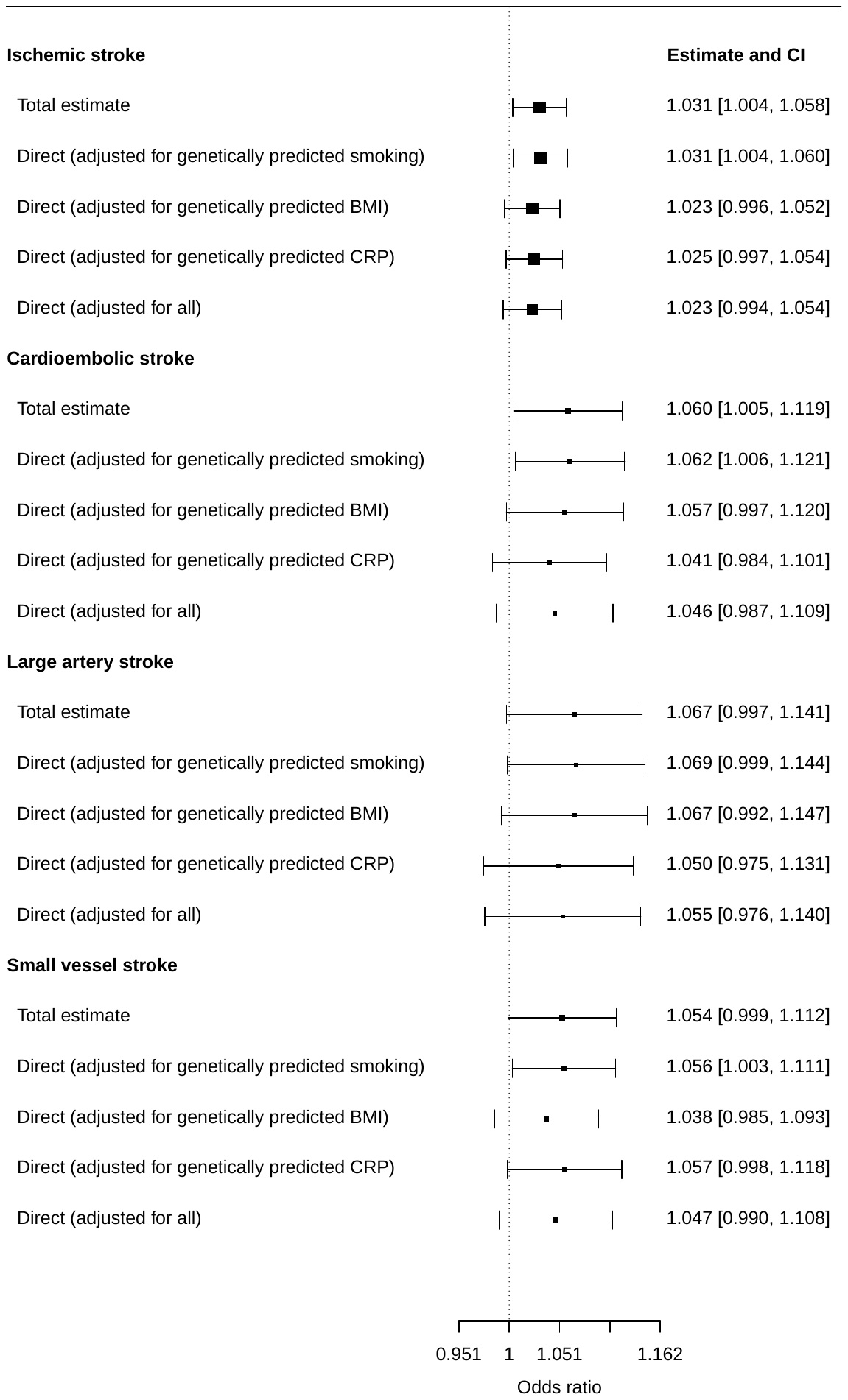

**Supplementary Figure 2**: Forest plot contrasting the Mendelian randomization estimates and confidence intervals (CI) from univariable Mendelian randomization (total estimate) and multivariable Mendelian randomization accounting for potential pleiotropic pathways (direct estimate). The total estimate of liability to critical Covid-19 on ischemic stroke outcomes was derived from a univariable (unadjusted) Mendelian randomization model; the direct estimate of Covid-19 on ischemic stroke outcomes was estimated in a multivariable Mendelian randomization model after adjusting for genetically predicted smoking intensity, body mass index (BMI), or C-reactive protein (CRP), and all three potential pleiotropic pathways jointly. Mendelian randomization estimates represent the odds ratio for ischemic stroke outcomes per unit increase in the log-odds ratio of liability to critical Covid-19.

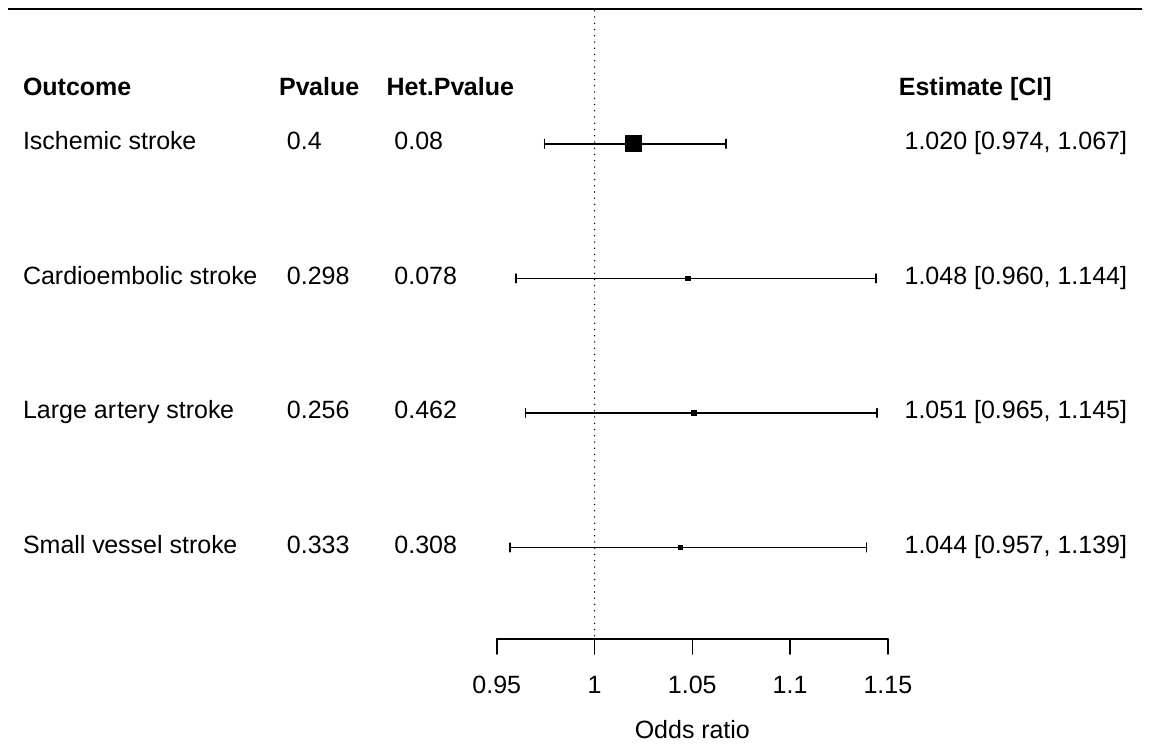

Supplementary Figure 3: Forest plot illustrating the Mendelian randomization estimates of liability to critical Covid-19 on stroke outcomes based on inverse-variance weighted Mendelian randomization using 9 genetic variants which were associated with liability to critical Covid-19 at genome-wide significance (*p*-value < *5×10^-8^*). Mendelian randomization estimates represent the odds ratio for ischemic stroke outcomes per unit increase in the log-odds ratio of critical Covid-19 liability. Additional columns include the *p-*value (*p*-value) of the Mendelian randomization estimate to be different from the null, represented by a dashed line at an odds ratio of 1, and the heterogeneity of the Mendelian randomization model measured by the heterogeneity *p-*value (Het. *p*-value) as well as the Mendelian randomization estimate and its 95% confidence interval (CI).

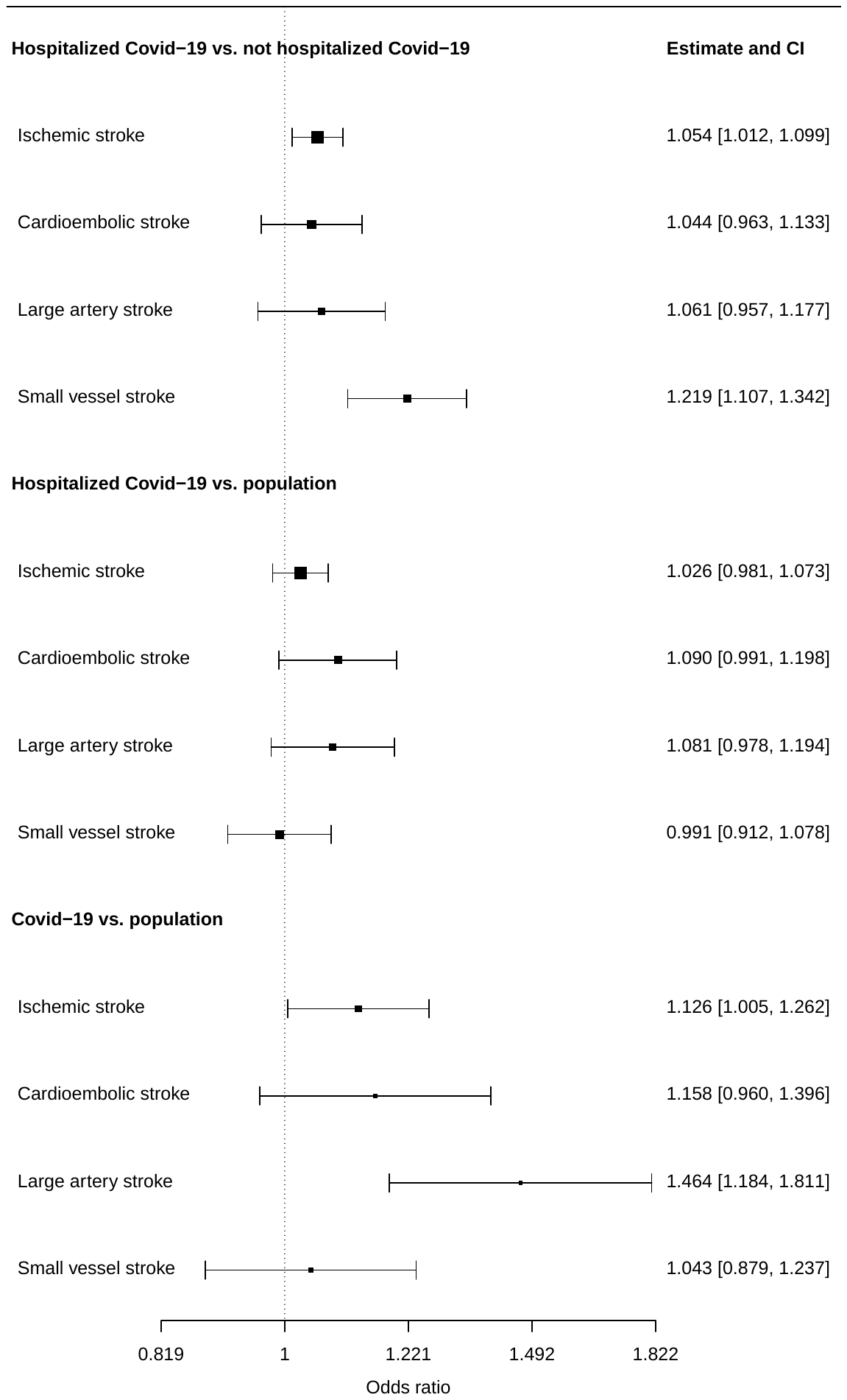

**Supplementary Figure 4**: Forest plot illustrating the inverse-variance weighted Mendelian randomization estimate and 95% confidence interval (CI) considering Covid-19 phenotypes as exposure for ischemic stroke outcomes. Covid-19 phenotypes were based on the definitions by the Covid-19 host genetics initiative. Mendelian randomization estimates represent the odds ratio for ischemic stroke outcomes per unit increase in the log-odds ratio of liability to the respective Covid-19 definition. Genetic variants which were associated with the Covid-19 definition were selected as instrumental variables at a *p-*value level equal to *5×10^-6^* or smaller.
